# Medicaid and the Type of Post-Acute Care Among Working-Age Survivors of Intensive Care: A Two-Setting Comparison of a Massachusetts Academic Medical Center and Texas Hospitalizations With ICU or Coronary Care Charges

**DOI:** 10.64898/2026.11.13.26368054

**Authors:** Mohammad Nasir Uddin

## Abstract

Insurance-related differences in post-acute care after critical illness have mostly been studied within single conditions or settings. We described Medicaid–private differences in discharge destination and length of stay among working-age adults in two non-equivalent settings: 20,511 patients whose first hospitalization included an ICU stay at one Massachusetts academic medical center (2008–2022), and 603,512 Texas hospitalizations with ICU or coronary care unit accommodation charges in 325 acute care hospitals (2017–2019). Adjusted for admission-time characteristics, Medicaid was associated in both settings with more skilled nursing facility discharge (5.2 and 4.3 percentage points), less inpatient rehabilitation (−2.0 and −2.6 points), and longer stays. Long-term acute care and total facility use differed in direction. Destination differences persisted with further adjustment and in a common 2017–2019 period; length-of-stay differences narrowed. Texas Medicaid rules limiting adult freestanding rehabilitation and inpatient days are consistent with, but cannot establish, a role for benefit design.

## Introduction

Survivors of critical illness often leave the hospital with new physical, cognitive, and mental health impairments (Needham et al., 2012). Where they go next — home, home with home health care, a skilled nursing facility (SNF), an inpatient rehabilitation facility (IRF), or a long-term acute care hospital (LTCH) — determines the setting and intensity of services they receive after discharge. IRFs provide intensive hospital-level rehabilitation, LTCHs extended hospital care for prolonged critical illness (Kahn, Benson, et al., 2010), and SNFs skilled nursing and therapy at lower intensity. Which setting a patient enters depends on clinical need and also on coverage, authorization, and provider acceptance.

Insurance has been associated with post-acute care in several populations. Among working-age stroke survivors, Medicaid was associated with more institutional post-acute care but, among institutional users, with less IRF and more SNF care (Skolarus et al., 2012), and a national stroke registry found Medicaid patients more likely than privately insured patients to be discharged to an SNF and less likely to an IRF (Medford-Davis et al., 2016). Among working-age stroke patients with Medicaid in 2010, IRF discharge was less common in states whose Medicaid programs did not cover IRF care, a group that included Texas (Skolarus et al., 2014). Among mechanically ventilated patients in Pennsylvania, Medicaid was associated with lower LTCH use than commercial insurance (Lane-Fall et al., 2012). Findings differ across conditions (Kane et al., 2014), and Medicaid expansion has been associated with more rehabilitation after injury (Zogg et al., 2019). In the database used here, two recent conference abstracts examined insurance and ICU length of stay among patients with stroke (Shah, 2025) and discharge to inpatient rehabilitation among ICU patients with ischemic stroke treated with thrombolysis (Shah & Strobel, 2026); both concern a single condition and a single outcome and have not been peer reviewed. Insurance may also be related to length of stay: authorization and bed availability are commonly documented reasons for delayed discharge (Sorensen et al., 2020), and among injured patients discharged to facilities, those enrolled in Medicaid retroactively during the admission had longer stays than those with established Medicaid coverage (Haddad et al., 2025). In Massachusetts, non-peer-reviewed hospital association and state task force reports describe payer authorization, including by commercial insurers, as a source of delayed transitions (Massachusetts Health & Hospital Association, 2025; Massachusetts TACPAC Task Force, 2025).

Two gaps remain. First, most studies concern a single diagnosis or a single post-acute setting, so they cannot show whether insurance is related to *whether* patients receive facility-based care, *which* facility they receive, or both. Second, single-site or single-state studies cannot show which associations recur across settings that differ in their data, populations, and Medicaid programs.

We therefore described Medicaid–private differences in the full distribution of discharge destinations and in length of stay among working-age survivors of intensive care in two non-equivalent settings: one academic medical center in Massachusetts, a state that expanded Medicaid, and acute care hospitals in Texas, a state that did not (Kaiser Family Foundation & Georgetown University Center for Children and Families, 2019). We asked which associations were similar across settings and which differed. We then compared the differences with documented Medicaid benefit rules in each state as a way to generate hypotheses, not to estimate policy effects.

### New Contribution

The study adds three things to prior work. First, it reports every discharge destination separately in a broad, all-diagnosis population of working-age survivors of intensive care, which shows that the most consistent insurance difference concerns the *type* of post-acute care (nursing facility rather than inpatient rehabilitation) more than the *amount* of facility-based care; a single facility composite would have obscured offsetting differences. Earlier analyses of the same database were limited to stroke and to one outcome (Shah, 2025; Shah & Strobel, 2026). Second, it pairs a clinically detailed single-center cohort with a large discharge dataset from 319 Texas hospitals analyzed within hospitals, so that the main associations do not rest on one institution’s case mix or referral network. Third, it documents the Texas Medicaid benefit rules for adult IRF care and inpatient days in force during the study years, including the programs to which they applied, and shows that the Texas pattern is consistent with them, extending stroke-specific evidence on IRF coverage (Skolarus et al., 2014) to critical illness and to LTCH care. The comparison across settings is descriptive and generates hypotheses about benefit design.

### Conceptual Framework

We used the behavioral model of health services use, in which use reflects need, predisposing characteristics, and enabling resources (Andersen, 1995). After critical illness, need is reflected in comorbidity, acute severity, and organ support; predisposing characteristics include age, sex, and race and ethnicity; and insurance is an enabling resource. We distinguish three ways insurance may be related to post-acute care. Through *benefit design*, a payer may not cover a setting for adults or may limit inpatient days. Through *administration*, prior authorization and network rules may delay or prevent placement. Through *provider acceptance*, facilities may decline a payer’s rates. Other enabling factors, such as caregiver support, housing, and substance use, are correlated with insurance and may independently limit discharge home.

These channels suggest patterns, not tests. Provider acceptance and social circumstances would be expected to produce less discharge home and more facility use with Medicaid in both settings. Benefit design would be expected to make the *type* of facility track a state’s rules, with less use of settings that its Medicaid program covers narrowly for adults and substitution toward settings it covers. Administrative and placement delays would be expected to lengthen hospital stays, particularly among patients discharged to facilities. Because insurance is recorded at admission and several clinical measures accrue during the stay and may themselves reflect care processes, the framework also orders covariates in time (Figure 1; Supplementary Table S2), which determined the estimand and the sequence of models.

**Figure 1.**
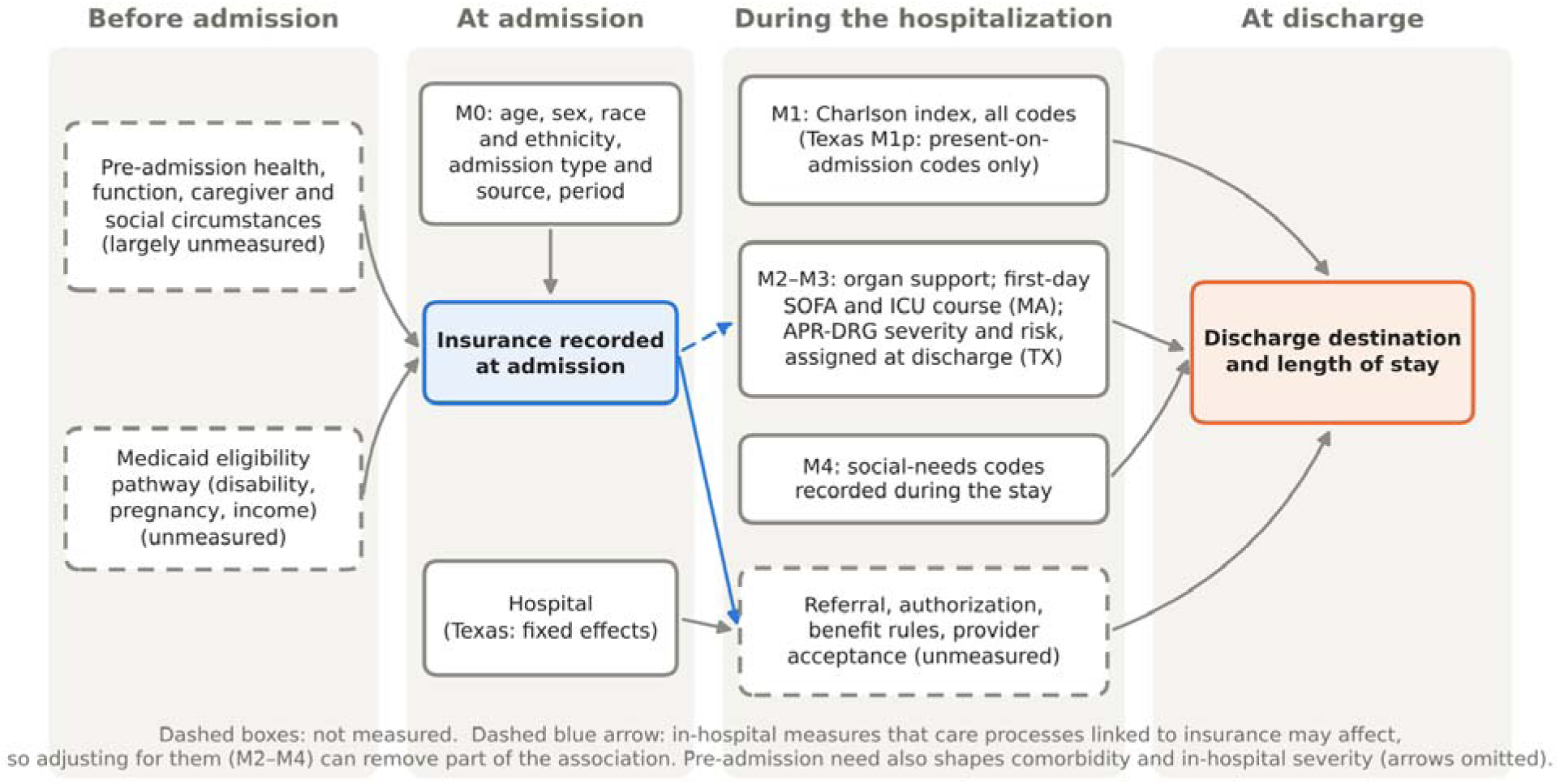
Temporal ordering of covariates relative to insurance and the outcomes. Note. Covariate sets M0–M4 are added in the order shown. Only M0 characteristics are clearly measured before insurance could affect care; later measures may lie on pathways from insurance to the outcomes (dashed blue arrow) or be coded differently by payer, so models that include them estimate conditional associations. Unmeasured factors (dashed boxes) can confound any model.

## Method

### Design and data sources

This was a retrospective, two-setting cohort study with separate data sources and analyses; the settings were not pooled. Setting 1 is a single academic medical center in Boston, Massachusetts. Setting 2 is acute care hospitals in Texas that reported to the state discharge data system. Reporting follows the STROBE and RECORD guidelines (von Elm et al., 2007; Benchimol et al., 2015); the checklist is in Supplementary Table S13.

#### Setting 1 (Massachusetts)

We used MIMIC-IV version 3.1 (Johnson, Bulgarelli, Shen, et al., 2023; Johnson, Bulgarelli, Pollard, et al., 2024), de-identified electronic health records for patients admitted to the emergency department or ICUs of the medical center from 2008 to 2022, accessed through PhysioNet (Pollard et al., 2026) under a credentialed data use agreement. Severity and treatment variables came from the MIMIC Code Repository derived tables (Johnson et al., 2018). For each patient we selected the first hospitalization that included an ICU stay, before applying other criteria; the unit of analysis is the patient.

#### Setting 2 (Texas)

We used the Texas Hospital Inpatient Discharge Public Use Data File (PUDF), which contains discharges reported by state-licensed hospitals; reporting exemptions for rural hospitals were repealed beginning with 2015 data (Texas Department of State Health Services, 2020, 2021). The primary period was 2017–2019; 2020 was analyzed separately, after removing outpatient records that the state identified in the second-quarter 2020 file. The PUDF has no patient identifier, so the unit of analysis is the hospitalization. Eligible stays were hospitalizations with ICU or coronary care unit (CCU) accommodation charges, recorded under revenue centers 020X and 021X (Texas Department of State Health Services, 2020). The ICU category also includes intermediate (step-down), burn, trauma, and psychiatric intensive care charges, which the file reports as a single amount, so stays with any of these charges enter the cohort and the Texas cohort is a billing-based construct rather than a validated clinical ICU cohort. We therefore report results separately for stays with ICU charges only and CCU charges only, and after excluding stays in mental health and substance use diagnostic categories and in psychiatric-designated hospitals. The specialty-unit field cannot serve as an independent check: the ICU or CCU was coded as the first specialty unit for 99% of eligible stays.

### Study population

In both settings we included adults aged 18–64 years who survived to discharge, were not discharged to hospice, had Medicaid or private insurance, and had a classifiable discharge destination; Setting 1 also required a valid interval between final ICU exit and discharge. Texas stays at hospitals with masked identifiers or at specialty hospitals without an acute care indicator were excluded. On Texas records with HIV or substance use diagnoses, the state reports age in coarsened groups (18–44 and 45–64 years) and masks sex; in the primary models these records keep their coarsened age group as its own category and an *unknown* sex category, and sensitivity analysis S9 excludes them. Sample flows are shown in Supplementary Figure S1 and, for Texas by payer, Supplementary Table S1.

### Exposure

In Setting 1 the exposure was admission-recorded insurance (Medicaid or private). In Setting 2 it was the expected primary payer on the claim: Medicaid, or private (commercial preferred provider, point-of-service, exclusive provider, indemnity, health maintenance organization, Blue Cross Blue Shield, or other commercial plans); Medicare Advantage claims were classified as Medicare (Supplementary Table S3). Both are administrative labels. Neither source identifies Medicaid eligibility group, managed care program or plan, or dual eligibility, and some Texas Medicaid managed care claims may be filed under commercial health maintenance organization codes.

### Outcomes

The author set the outcomes and their priority before the Texas analyses were run; the hierarchy was not prospectively registered. The primary outcomes were discharge to an SNF or nursing facility, discharge to an IRF, and hospital length of stay (LOS). Secondary outcomes were discharge home, with home health care, to an LTCH, to other destinations (against medical advice, transfer to another acute or psychiatric hospital, custodial care, and other), and to any post-acute facility (SNF, IRF, or LTCH). Destinations are harmonized administrative categories built from different source fields (Supplementary Table S3), not necessarily identical clinical settings; the Texas nursing facility category combines Medicare-certified SNFs and Medicaid-only nursing facilities, which we also report separately. LOS was measured in both settings as the whole hospital stay in calendar days (discharge date minus admission date, minimum 1), the definition available in Texas. In Setting 1 only, we also measured days from final ICU exit to hospital discharge (post-ICU days), treating negative intervals of up to 4 hours as zero. Neither LOS measure identifies time medically ready for discharge, placement delay, or avoidable days.

### Covariates and estimand

Admission-time covariates in Setting 1 were age (linear and quadratic), sex, race and ethnicity, admission type, and admission period; in Setting 2 they were age group, sex, race, ethnicity, admission type and source, emergency department use, and discharge quarter. Comorbidity was measured with the Charlson index without age points (Charlson et al., 1987; Quan et al., 2005). Its timing differs by source: MIMIC-IV diagnosis codes are assigned at discharge and carry no present-on-admission indicator, so the Setting 1 index may include conditions that arose during the stay; in Setting 2 we computed the index from all diagnosis codes and, as a check, only from codes flagged as present on admission or exempt from that reporting. Organ support and severity measures were, in Setting 1, the first-day Sequential Organ Failure Assessment score (Vincent et al., 1996), invasive ventilation, vasopressor use, ICU length of stay, and first ICU type, and in Setting 2, invasive ventilation, ventilation for 96 hours or more, and the All Patient Refined Diagnosis Related Group (APR-DRG) severity of illness, risk of mortality, and major diagnostic category, which are assigned at discharge from all of the stay’s codes. Diagnosis-coded social-needs proxies (housing or economic circumstances, substance use disorder, serious mental illness; Supplementary Table S3) are coded during the encounter and do not measure baseline social need. Race and ethnicity, recorded administratively, were treated as social constructs.

#### Estimand

For each outcome, the target quantity is the difference between Medicaid and privately insured patients (Setting 1) or stays (Setting 2) in the probability of each destination, or in mean LOS, standardized to the covariate distribution of the analytic sample — in Texas, among stays in the same hospital. It is a descriptive association, not the causal effect of insurance. Figure 1 orders the covariates in time. Only the admission-time characteristics are clearly measured before insurance could influence care, so the admission-time model (M0) is the primary descriptive comparison. We then added, in temporal order, comorbidity (M1); organ support (M2); in Texas, discharge-assigned severity of illness, risk of mortality, and major diagnostic category, and in Massachusetts, first-day severity and the ICU course (M3); and diagnosis-coded social-needs proxies (M4). Because later covariates may lie on pathways from insurance to the outcome or be coded differently by payer, M1–M4 are conditional, post-admission-adjusted associations and do not show whether severity explains a payer difference. We report M3, the covariate set of the earlier single-center analysis, alongside M0 throughout.

### Statistical analysis

#### Setting 1

Destination was modeled with multinomial logistic regression, and adjusted probabilities were obtained by marginal standardization (g-computation): each patient’s probabilities were predicted with insurance set to Medicaid and to private and then averaged over the cohort (Muller & MacLehose, 2014). We report adjusted probability differences and ratios, with 95% confidence intervals (CIs) from 500 patient-level bootstrap replicates (fixed seed; all replicates converged for the primary models). Differences were computed from unrounded probabilities. Post-ICU days were modeled with a two-part model (Deb & Norton, 2018). Robustness analyses included inverse probability weighting with stabilized, truncated weights (Austin & Stuart, 2015), with diagnostics in Supplementary Table S10, and the sequential models fitted as linear probability models with heteroskedasticity-robust errors.

#### Setting 2

Each destination was modeled separately with a logistic model that included a fixed effect for every hospital, so that comparisons are within hospitals and every predicted probability lies between 0 and 1. Adjusted probabilities were obtained by marginal standardization over all analytic stays (Muller & MacLehose, 2014), and we report adjusted probability differences and ratios with delta-method 95% CIs from a hospital-clustered sandwich variance; hospitals in which no stay had a given outcome contribute equal predicted probabilities under both payers. Because separate binary models need not sum to 1 across destinations, we also report a multinomial model without hospital fixed effects (Supplementary Table S6). We assessed calibration by deciles of predicted probability overall and by payer, the distribution of fitted probabilities by payer and across hospitals, and influence by approximate leave-one-hospital-out estimates and by excluding the 10 largest hospitals. Linear models with hospital and discharge-quarter fixed effects and hospital-clustered (CR1) standard errors were used for LOS, for comparison with the logistic destination models, and for the sensitivity analyses. We also report the hospitals that identify the within-hospital contrast and estimates restricted to hospitals with at least 11 stays of each payer and Medicaid shares of 5%–95%.

#### Sensitivity to unmeasured confounding

For SNF, IRF, and LTCH discharge we computed E-values (VanderWeele & Ding, 2017) for the adjusted probability ratio (Medicaid vs private, private as reference; ratios below 1 were inverted) and for the confidence limit closest to 1, from the Massachusetts multinomial model and the Texas fixed-effects logistic model; Texas conditional risk ratios from a Poisson model with hospital fixed effects are shown for comparison (Supplementary Table S20). An E-value is the minimum strength of association, on the risk ratio scale, that an unmeasured confounder would need with both insurance and the outcome, conditional on the measured covariates, to explain away the estimate; it does not address selection, misclassification of exposure or outcome, or model misspecification.

#### Missing data

In Setting 1, no covariate values were missing in the analytic cohort; 265 patients without a recorded discharge destination were excluded (Supplementary Figure S1). In Setting 2, no values were imputed: unknown race, ethnicity, admission type, and admission source were modeled as explicit unknown categories, ungroupable APR-DRG records as their own severity and risk category, coarsened age as its own age category, and masked sex as unknown; fewer than 11 stays without a length of stay were excluded. Missing and masked values by payer are shown in Supplementary Table S9.

Texas sensitivity analyses repeated the linear model (S1) excluding discharges against medical advice and transfers to acute hospitals; (S2) restricting to stays with ICU or CCU as the first coded specialty unit (99% of stays, so not a meaningfully stricter definition); (S3) restricting to invasively ventilated stays, a selected high-acuity population rather than a validation sample; (S4) adding the social-needs proxies; (S5) in 2020 and in 2017–2020 combined; (S6) without APR-DRG covariates; (S7) excluding privately insured stays with Medicaid as a secondary payer; (S8) excluding transfers in from other facilities; (S9) excluding records with coarsened age and masked sex; (S10) trimming LOS at the 99th percentile; and (S11) excluding pregnancy and childbirth stays. We also estimated models in teaching and non-teaching hospitals and, as a secondary analysis, among adults aged 65 years or older comparing Medicaid with Medicare as the primary payer. Analyses other than the three primary outcomes are secondary, and we emphasize direction, magnitude, and consistency rather than P values.

#### Comparison of settings

We re-estimated the Setting 1 associations using the Texas outcome definitions (linear probability models and whole-stay LOS) with a covariate set restricted to measures available in both sources (age group, sex, race and ethnicity, admission type, Charlson index, invasive ventilation, and period), for all patients and for invasively ventilated patients. To align calendar time, we also repeated the Setting 1 standardized models (M0 and M3) among patients whose MIMIC-IV anchor year group was 2017–2019 and compared them with the Texas 2017–2019 estimates; because MIMIC-IV shifts dates and reports only a 3-year anchor year group for each patient, this subset approximates admissions in those years. Between-setting differences have variance equal to the sum of the two squared standard errors, each recovered from its 95% CI as (upper − lower)/3.92 (robust or bootstrap in Setting 1; hospital-clustered in Texas), treating the samples as independent. These contrasts describe differences between data sources that also differ in unit of analysis, hospital mix, covariates, and measurement; they do not isolate state policy.

#### Benefit rules

We abstracted adult Medicaid coverage rules for IRF care, LTCH care, and inpatient day and spending limits, with the programs and age groups to which each applied, from state provider manuals, regulations, and state reports dated within or near each setting’s study years (Supplementary Table S12). These rules were not linked to individual patients, because neither data source identifies eligibility group, managed care program, or plan, so the payer field does not identify exposure to any rule.

Analyses used Python 3.12 with fixed random seeds; Supplementary Table S21 lists software versions and maps every estimate to the script and output that produced it. Aggregate output suppressed cells with fewer than 11 patients or stays. Texas data were analyzed on a local computer in accordance with the state data use agreement; the conclusions are the author’s and not those of the Texas Department of State Health Services. Analysis code is publicly available (Data availability). Generative artificial intelligence tools assisted with code, text, and literature searching; the author ran and verified all analyses, and no patient-level data were provided to these tools.

### Ethics

Both data sources are de-identified. MIMIC-IV was approved by the institutional review boards of the source hospital and the Massachusetts Institute of Technology with a waiver of consent; the Texas PUDF is a public use file released under a data use agreement. Because the study used only de-identified data, it did not constitute human subjects research requiring additional review.

## Results

### Study samples

In Setting 1, 20,511 patients aged 18–64 years met the criteria, of whom 6,934 (33.8%) had Medicaid. In Setting 2, of 9,339,506 discharges in 2017–2019, 2,479,142 were adult stays with ICU or CCU charges and 1,222,806 were among patients aged 18–64 years; of these, 645,595 had Medicaid or private insurance, most exclusions being stays with Medicare (disability-related), self-pay, or charity coverage. Of these 645,595 stays, we excluded 22,364 ending in death, 10,618 discharged to hospice, 119 with the patient still in hospital, 19 with an invalid discharge status, 318 at hospitals with masked identifiers, 8,644 at specialty hospitals without an acute care indicator, and fewer than 11 without a length of stay, accounting for all 42,083 excluded stays (Supplementary Table S1 gives each step by payer). The analytic sample included 603,512 stays in 325 hospitals, of which 151,862 (25.2%) were covered by Medicaid; 319 hospitals had more than one stay and contribute to within-hospital estimates. Within-hospital contrasts were identified by 302 hospitals that treated both payer groups, which accounted for 99.7% of stays; 266 hospitals had at least 11 stays of each payer (99.3% of stays), and the hospital Medicaid share had a median of 24.2% (interquartile range, 11.7%–38.1%). Compared with privately insured patients in Texas, Medicaid patients were younger (aged 55–64 years, 26.9% vs 38.7%), more often Black (26.1% vs 14.1%) or Hispanic (36.0% vs 23.9%), more often admitted as emergencies (76.8% vs 64.0%), more often had major or extreme severity of illness (59.0% vs 47.7%) and invasive ventilation (10.0% vs 7.0%), and more often had coded substance use disorder (19.3% vs 10.9%) or serious mental illness (11.3% vs 3.3%; Table 1). The same differences in age, race and ethnicity, admission type, and social-needs codes were present in Massachusetts.

**Table 1.** Characteristics of working-age survivors of intensive care, by insurance and setting.

| Characteristic | Massachusetts: Medicaid (n = 6,934 patients) | Massachusetts: private (n = 13,577) | SMD | Texas: Medicaid (n = 151,862 stays) | Texas: private (n = 451,650) | SMD |
|---|---|---|---|---|---|---|
| **Age** |  |  |  |  |  |  |
| Mean (SD), years | 47.0 (12.3) | 49.7 (12.4) | −0.22 | — | — | — |
| 18–34 years, % | — | — | — | 22.1 | 13.9 | 0.21 |
| 35–44 years, % | — | — | — | 12.4 | 13.1 | −0.02 |
| 45–54 years, % | — | — | — | 17.4 | 22.7 | −0.13 |
| 55–64 years, % | — | — | — | 26.9 | 38.7 | −0.25 |
| Masked age group (HIV or substance use record), % | — | — | — | 21.1 | 11.6 | 0.26 |
| Female sex, % | 44.3 | 40.2 | 0.08 | 47.7 | 44.3 | 0.07 |
| Sex masked or unknown, % | 0 | 0 | — | 21.8 | 12.1 | 0.26 |
| **Race and ethnicity, %** |  |  |  |  |  |  |
| White | 50.8 | 67.8 | −0.35 | 56.1 | 68.5 | −0.26 |
| Black | 15.2 | 8.2 | 0.22 | 26.1 | 14.1 | 0.30 |
| Asian or Pacific Islander | 3.8 | 3.7 | 0.01 | 1.1 | 2.2 | −0.09 |
| Hispanic or Latino ethnicity | 9.2 | 3.0 | 0.26 | 36.0 | 23.9 | 0.27 |
| Other or unknown race | 21.0 | 17.3 | 0.09 | 16.7 | 15.2 | 0.04 |
| **Admission type, %** |  |  |  |  |  |  |
| Emergency | 55.3 | 50.1 | 0.10 | 76.8 | 64.0 | 0.28 |
| Urgent | 16.3 | 15.7 | 0.02 | 10.3 | 14.4 | −0.12 |
| Elective | 11.8 | 21.4 | −0.26 | 11.4 | 20.1 | −0.24 |
| Trauma, observation, or other | 16.5 | 12.8 | 0.10 | 1.6 | 1.5 | 0.01 |
| Transfer from another facility, % | — | — | — | 8.0 | 9.6 | −0.06 |
| **Severity and treatment** |  |  |  |  |  |  |
| First-day SOFA score, median (IQR) | 3 (1–5) | 2 (1–5) | — | — | — | — |
| APR-DRG severity of illness major or extreme, % | — | — | — | 59.0 | 47.7 | 0.23 |
| APR-DRG risk of mortality major or extreme, % | — | — | — | 40.7 | 31.1 | 0.20 |
| Charlson index excluding age, median (IQR) | 1 (0–3) | 1 (0–3) | — | — | — | — |
| Charlson index excluding age ≥ 3, % | — | — | — | 41.7 | 29.5 | 0.26 |
| Invasive mechanical ventilation, % | 39.6 | 37.7 | 0.04 | 10.0 | 7.0 | 0.11 |
| Ventilation ≥ 96 hours, % | — | — | — | 3.6 | 2.3 | 0.08 |
| Vasopressor use, % | 22.6 | 24.8 | −0.05 | — | — | — |
| **Diagnosis-coded social needs, %** |  |  |  |  |  |  |
| Substance use disorder | 35.1 | 14.0 | 0.51 | 19.3 | 10.9 | 0.24 |
| Serious mental illness | 8.9 | 2.4 | 0.28 | 11.3 | 3.3 | 0.31 |
| Housing or economic circumstances | 4.6 | 0.2 | 0.29 | 2.0 | 0.3 | 0.16 |
| **Hospital** |  |  |  |  |  |  |
| Teaching hospital, % | 100 | 100 | — | 35.9 | 33.5 | 0.05 |
| Hospital length of stay, days, mean (SD) | — | — | — | 6.97 (11.53) | 5.59 (7.11) | — |

Values are percentages unless stated. SMD, standardized mean difference (Medicaid – private) for the indicator in each row; values of 0.1 or more in absolute value indicate imbalance. Massachusetts: first hospitalization that included an ICU stay, per patient, at one academic medical center, 2008–2022. Texas: hospitalizations with ICU or coronary care unit accommodation charges in reporting acute care hospitals, 2017–2019; patients may contribute more than one stay. Measures not available in a data source are shown as —. Texas records with HIV or substance use diagnoses carry coarsened age groups and masked sex, shown in separate rows. Abbreviations: APR-DRG, All Patient Refined Diagnosis Related Group; IQR, interquartile range; SOFA, Sequential Organ Failure Assessment.

### Discharge destination

In Setting 1, adjusted for admission-time characteristics (M0), Medicaid was associated with a higher probability of SNF discharge (adjusted probability difference, 5.2 percentage points [pp]; 95% CI, 4.4 to 6.0) and of LTCH discharge (2.1 pp; 1.5 to 2.7), and with lower probabilities of discharge home (−6.3 pp; −7.8 to −4.9), with home health (−3.5 pp; −4.8 to −2.2), and to an IRF (−2.0 pp; −2.7 to −1.2; Table 2). With the full covariate set (M3), the corresponding differences were SNF 4.6 pp (3.8 to 5.4; adjusted probabilities 8.8% vs 4.3%), LTCH 1.4 pp (0.8 to 1.9), home −4.8 pp (−6.1 to −3.5), home health −3.3 pp (−4.5 to −2.0), and IRF −1.7 pp (−2.5 to −0.9).

**Table 2.**
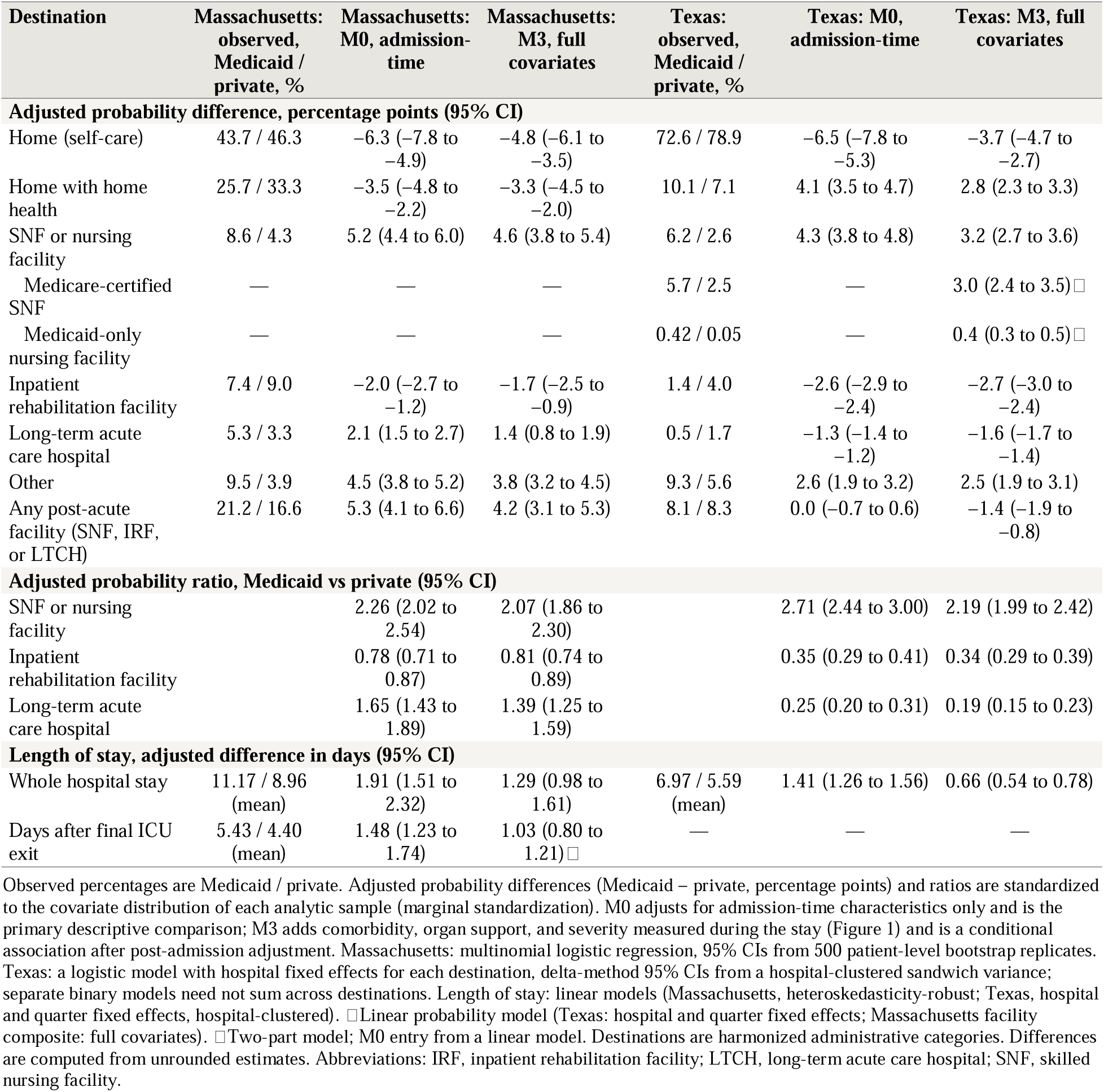
Discharge destination and length of stay, Medicaid versus private insurance, by setting and adjustment set.

| Destination | Massachusetts: observed, Medicaid / private, % | Massachusetts: M0, admission-time | Massachusetts: M3, full covariates | Texas: observed, Medicaid / private, % | Texas: M0, admission-time | Texas: M3, full covariates |
|---|---|---|---|---|---|---|
| **Adjusted probability difference, percentage points (95% CI)** |  |  |  |  |  |  |
| Home (self-care) | 43.7 / 46.3 | −6.3 (−7.8 to −4.9) | −4.8 (−6.1 to −3.5) | 72.6 / 78.9 | −6.5 (−7.8 to −5.3) | −3.7 (−4.7 to −2.7) |
| Home with home health | 25.7 / 33.3 | −3.5 (−4.8 to −2.2) | −3.3 (−4.5 to −2.0) | 10.1 / 7.1 | 4.1 (3.5 to 4.7) | 2.8 (2.3 to 3.3) |
| SNF or nursing facility | 8.6 / 4.3 | 5.2 (4.4 to 6.0) | 4.6 (3.8 to 5.4) | 6.2 / 2.6 | 4.3 (3.8 to 4.8) | 3.2 (2.7 to 3.6) |
| Medicare-certified SNF | — | — | — | 5.7 / 2.5 | — | 3.0 (2.4 to 3.5)□ |
| Medicaid-only nursing facility | — | — | — | 0.42 / 0.05 | — | 0.4 (0.3 to 0.5)□ |
| Inpatient rehabilitation facility | 7.4 / 9.0 | −2.0 (−2.7 to −1.2) | −1.7 (−2.5 to −0.9) | 1.4 / 4.0 | −2.6 (−2.9 to −2.4) | −2.7 (−3.0 to −2.4) |
| Long-term acute care hospital | 5.3 / 3.3 | 2.1 (1.5 to 2.7) | 1.4 (0.8 to 1.9) | 0.5 / 1.7 | −1.3 (−1.4 to −1.2) | −1.6 (−1.7 to −1.4) |
| Other | 9.5 / 3.9 | 4.5 (3.8 to 5.2) | 3.8 (3.2 to 4.5) | 9.3 / 5.6 | 2.6 (1.9 to 3.2) | 2.5 (1.9 to 3.1) |
| Any post-acute facility (SNF, IRF, or LTCH) | 21.2 / 16.6 | 5.3 (4.1 to 6.6) | 4.2 (3.1 to 5.3) | 8.1 / 8.3 | 0.0 (−0.7 to 0.6) | −1.4 (−1.9 to −0.8) |
| **Adjusted probability ratio, Medicaid vs private (95% CI)** |  |  |  |  |  |  |
| SNF or nursing facility |  | 2.26 (2.02 to 2.54) | 2.07 (1.86 to 2.30) |  | 2.71 (2.44 to 3.00) | 2.19 (1.99 to 2.42) |
| Inpatient rehabilitation facility |  | 0.78 (0.71 to 0.87) | 0.81 (0.74 to 0.89) |  | 0.35 (0.29 to 0.41) | 0.34 (0.29 to 0.39) |
| Long-term acute care hospital |  | 1.65 (1.43 to 1.89) | 1.39 (1.25 to 1.59) |  | 0.25 (0.20 to 0.31) | 0.19 (0.15 to 0.23) |
| **Length of stay, adjusted difference in days (95% CI)** |  |  |  |  |  |  |
| Whole hospital stay | 11.17 / 8.96 (mean) | 1.91 (1.51 to 2.32) | 1.29 (0.98 to 1.61) | 6.97 / 5.59 (mean) | 1.41 (1.26 to 1.56) | 0.66 (0.54 to 0.78) |
| Days after final ICU exit | 5.43 / 4.40 (mean) | 1.48 (1.23 to 1.74) | 1.03 (0.80 to 1.21)□ | — | — | — |

Observed percentages are Medicaid / private. Adjusted probability differences (Medicaid – private, percentage points) and ratios are standardized to the covariate distribution of each analytic sample (marginal standardization). M0 adjusts for admission-time characteristics only and is the primary descriptive comparison; M3 adds comorbidity, organ support, and severity measured during the stay (Figure 1) and is a conditional association after post-admission adjustment. Massachusetts: multinomial logistic regression, 95% CIs from 500 patient-level bootstrap replicates. Texas: a logistic model with hospital fixed effects for each destination, delta-method 95% CIs from a hospital-clustered sandwich variance; separate binary models need not sum across destinations. Length of stay: linear models (Massachusetts, heteroskedasticity-robust; Texas, hospital and quarter fixed effects, hospital-clustered). □Linear probability model (Texas: hospital and quarter fixed effects; Massachusetts facility composite: full covariates). □Two-part model; M0 entry from a linear model. Destinations are harmonized administrative categories. Differences are computed from unrounded estimates. Abbreviations: IRF, inpatient rehabilitation facility; LTCH, long-term acute care hospital; SNF, skilled nursing facility.

In Setting 2, crude SNF discharge was more than twice as common with Medicaid (6.2% vs 2.6%), whereas IRF (1.4% vs 4.0%) and LTCH discharge (0.5% vs 1.7%) were much less common (Table 2). Within hospitals and adjusted for admission-time characteristics (M0), Medicaid was associated with more SNF or nursing facility discharge (4.3 pp; 95% CI, 3.8 to 4.8), more discharge with home health (4.1 pp; 3.5 to 4.7), less IRF (−2.6 pp; −2.9 to −2.4) and LTCH discharge (−1.3 pp; −1.4 to −1.2), and less discharge home (−6.5 pp; −7.8 to −5.3). With the full covariate set (M3), the differences were SNF 3.2 (2.7 to 3.6), IRF −2.7 (−3.0 to −2.4), and LTCH −1.6 (−1.7 to −1.4) pp. Most of the SNF difference was in Medicare-certified SNFs (3.0 pp in the linear M3 model; 2.4 to 3.5). Because the IRF and LTCH differences offset the SNF difference, total facility discharge differed little by payer (M0, 0.0 pp; −0.7 to 0.6; M3, −1.4 pp; −1.9 to −0.8). All fitted probabilities from the fixed-effects logistic model lie between 0 and 1, whereas the linear models produced fitted values below zero for 27.6% of stays for IRF and 31.9% for LTCH. The two models nonetheless gave similar adjusted probability differences (M3 linear model: SNF 3.4, IRF −3.0, LTCH −2.0 pp), and the logistic model’s predicted probabilities tracked observed rates across deciles of predicted risk in both payer groups (Supplementary Tables S17–S18).

### Length of stay

Medicaid was associated with longer stays in both settings. Adjusted for admission-time characteristics, whole-stay LOS was 1.91 days longer in Setting 1 and 1.41 days longer within Texas hospitals; with the full covariate set, the differences were 1.29 days (95% CI, 0.98 to 1.61) and 0.66 days (0.54 to 0.78; ratio of geometric means 1.04). In Setting 1, Medicaid was also associated with 1.03 more post-ICU days in the two-part model (95% CI, 0.80 to 1.21). In both settings the LOS difference was larger among patients discharged to a post-acute facility than among those discharged home (Setting 1, 2.30 vs 0.65 post-ICU days; Setting 2, 2.25 [1.42 to 3.08] vs 0.63 [0.54 to 0.72] whole-stay days); because destination may itself depend on insurance, this stratified comparison is descriptive.

### Sequential adjustment

In the linear models, the destination associations changed modestly as covariates were added in temporal order (the SNF difference narrowed and the IRF difference widened slightly), whereas the LOS associations narrowed more (Table 5 and Supplementary Table S5). In Texas, the SNF difference was 4.1 pp with admission-time characteristics only (M0), 3.4 pp in M3, and 3.1 pp with social-needs proxies (M4), and the IRF difference was −2.6 pp in M0 and −3.0 pp in M3; the total facility difference was near zero in M0 (0.1 pp) and −1.6 pp in M3. In Massachusetts, the SNF difference was 5.0 pp in M0 and 4.4 pp in M3, and the IRF difference −2.0 and −1.9 pp. The LOS difference fell from 1.41 to 0.66 days in Texas and from 1.91 to 1.29 days in Massachusetts (post-ICU days, 1.48 to 1.12), mostly when comorbidity, organ support, and severity were added, so part of the longer stay with Medicaid reflects measured severity. In Texas, present-on-admission indicators were reported on 99.8% of Medicaid and 99.8% of private stays, and computing the Charlson index only from present-on-admission codes changed the estimates little (fixed-effects logistic model, M1p vs M1: SNF 3.7 vs 3.6 pp; IRF −2.9 vs −2.9 pp; M3p: SNF 3.2, IRF −2.7 pp; Supplementary Table S17). These comparisons show how the estimates depend on the covariate set; small changes across models do not show that unmeasured need, severity, or access mechanisms are unimportant.

### Robustness

Texas estimates were similar across sensitivity analyses (Table 4). In the linear models, the SNF difference ranged from 3.1 to 3.7 pp and the IRF difference from −3.2 to −2.8 pp across S1, S2, and S4–S11, and the LOS difference from 0.6 to 0.8 days (0.2 days with LOS trimmed at the 99th percentile). In the fixed-effects logistic model (M3), the SNF and IRF differences were 3.2 and −2.7 pp among stays with ICU charges only, 2.9 and −1.5 pp among stays with CCU charges only, and 3.2 and −2.7 pp after excluding mental health and substance use diagnostic categories and psychiatric-designated hospitals (Supplementary Table S19). Without hospital fixed effects, the SNF difference was 3.0 pp and the LOS difference 0.79 days, close to the within-hospital estimates (Supplementary Table S4). No single hospital drove the results: leaving out one hospital at a time, the linear-model SNF difference ranged from 3.21 to 3.43 pp and the IRF difference from −3.06 to −2.95 pp; restricting to hospitals with good payer overlap gave SNF 3.4 and IRF −3.0 pp, and excluding the 10 largest hospitals gave fixed-effects logistic differences of 3.3 and −2.7 pp (Supplementary Tables S7 and S18). Among invasively ventilated stays (S3; 46,681 stays in 267 hospitals), a selected high-acuity population, the differences were larger: SNF 8.7 pp (7.4 to 10.1), IRF −8.9 pp (−9.9 to −7.9), total facility discharge −11.0 pp (−12.6 to −9.4), and LOS 3.5 days (2.8 to 4.3). For the adjusted probability ratios (Medicaid vs private) in M3, E-values (point estimate and confidence limit) were, in Massachusetts, 3.56 (confidence limit, 3.12) for SNF (ratio 2.07; 1.86 to 2.30), 1.77 (confidence limit, 1.49) for IRF (0.81; 0.74 to 0.89), and 2.13 (confidence limit, 1.81) for LTCH (1.39; 1.25 to 1.59); in Texas, 3.81 (confidence limit, 3.40) for SNF (2.19; 1.99 to 2.42), 5.39 (confidence limit, 4.55) for IRF (0.34; 0.29 to 0.39), and 10.16 (confidence limit, 8.05) for LTCH (0.19; 0.15 to 0.23; Supplementary Table S20). These values describe sensitivity to unmeasured confounding under the E-value assumptions, conditional on the measured covariates; they do not show that residual confounding is absent. Results among adults aged 65 years or older, a secondary and atypical comparison of Medicaid-primary with Medicare-primary stays, are in Supplementary Table S11.

### Comparison of the two settings

With harmonized definitions (Table 3 and Figure 2), both settings showed more SNF discharge, less IRF discharge, less discharge home, and longer whole-stay LOS with Medicaid. The SNF difference was somewhat larger in Massachusetts (4.4 vs 3.4 pp; difference between settings 1.1; 95% CI, 0.1 to 2.0 pp), and the IRF differences were similar (−2.6 vs −3.0 pp; 0.4; 95% CI, −0.5 to 1.3 pp). The settings differed in total facility discharge (3.3 vs −1.6 pp; 4.9; 95% CI, 3.6 to 6.2 pp), reflecting the opposite LTCH associations, and the LOS difference was larger in Massachusetts (1.25 vs 0.66 days; 0.59; 95% CI, 0.19 to 0.99 days). Among ventilated patients, the SNF and IRF differences were larger in Texas (SNF 8.7 vs 5.3 pp, difference −3.5; 95% CI, −5.4 to −1.5 pp; IRF −8.9 vs −5.2 pp, 3.6; 95% CI, 1.8 to 5.5 pp), as was the LOS difference (3.5 vs 1.6 days). Restricting Setting 1 to the 2017–2019 anchor year group (4,018 patients, 1,394 with Medicaid) gave admission-time SNF and IRF differences of 5.2 pp; 3.5 to 7.0 and −1.6 pp; −3.4 to 0.1, compared with 4.3 and −2.6 pp in Texas (differences between settings, 0.9 pp; −0.9 to 2.8 and 1.1 pp; −0.7 to 2.8), and an LTCH difference of 3.1 pp; 1.8 to 4.5, compared with −1.3 pp in Texas (Table 3). The subset estimates were less precise (the IRF interval included zero; 459 and 457 of 500 bootstrap replicates converged for M0 and M3), and the whole-stay LOS difference in the subset was larger than in the full cohort (M3, 2.24 days; 1.31 to 3.17).

**Figure 2.**
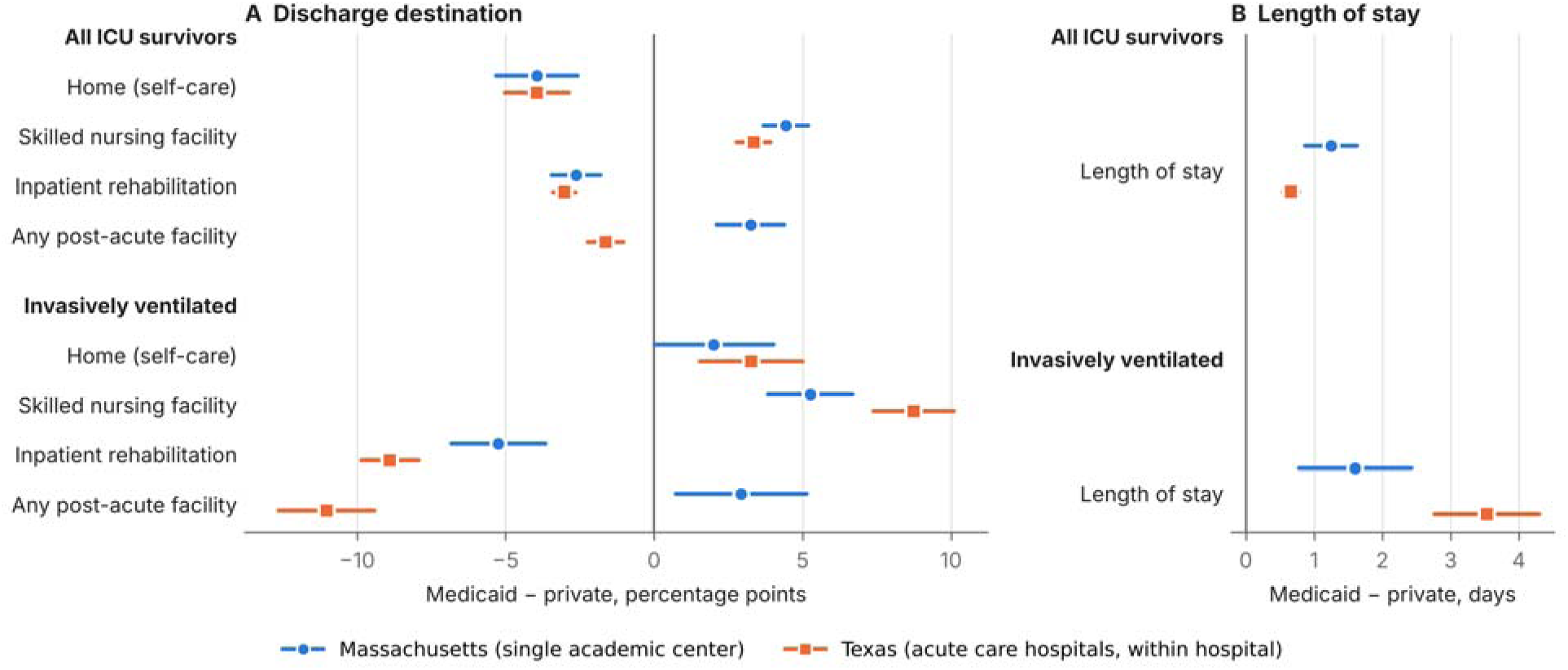
Medicaid − private differences in discharge destination and length of stay in Massachusetts and Texas, harmonized definitions. Note. Points are adjusted probability differences (length of stay: differences in days) and lines are 95% CIs. Massachusetts (circles): one academic medical center, linear probability models with Texas-comparable covariates. Texas (squares): linear probability models with hospital and quarter fixed effects in reporting acute care hospitals; ventilated estimates are sensitivity analysis S3. Differences between settings are descriptive (Table 3). Length of stay is the whole hospital stay in days.

**Table 3.** Harmonized and common-period comparison of the two settings.

| **Cohort and outcome** | **Massachusetts: full covariates (panels A–B) or common period (panel C)** | **Massachusetts: Texas-comparable covariates** | **Texas** | **Difference, Massachusetts – Texas** |
|---|---|---|---|---|
| **A. All patients or stays, linear probability models (Massachusetts n = 20,511; Texas n = 603,512)** |  |  |  |  |
| Home (self-care) | −5.0 (−6.3 to −3.6) | −4.0 (−5.3 to −2.6) | −3.9 (−5.0 to −2.9) | 0.0 (−1.8 to 1.7) |
| SNF or nursing facility | 4.4 (3.7 to 5.2) | 4.4 (3.7 to 5.2) | 3.4 (2.8 to 3.9) | 1.1 (0.1 to 2.0) |
| Inpatient rehabilitation facility | −1.9 (−2.7 to −1.1) | −2.6 (−3.4 to −1.8) | −3.0 (−3.4 to −2.6) | 0.4 (−0.5 to 1.3) |
| Any post-acute facility | 4.0 (3.0 to 5.1) | 3.3 (2.1 to 4.4) | −1.6 (−2.2 to −1.0) | 4.9 (3.6 to 6.2) |
| Whole-stay length of stay, days | 1.29 (0.98 to 1.61) | 1.25 (0.87 to 1.63) | 0.66 (0.54 to 0.78) | 0.59 (0.19 to 0.99) |
| **B. Invasively ventilated, linear probability models (Massachusetts n = 7,861; Texas n = 46,681)** |  |  |  |  |
| Home (self-care) | −0.5 (−2.4 to 1.5) | 2.0 (0.0 to 4.0) | 3.3 (1.5 to 5.0) | −1.3 (−3.9 to 1.4) |
| SNF or nursing facility | 5.5 (4.0 to 6.9) | 5.3 (3.8 to 6.7) | 8.7 (7.4 to 10.1) | −3.5 (−5.4 to −1.5) |
| Inpatient rehabilitation facility | −4.5 (−6.0 to −3.0) | −5.2 (−6.8 to −3.7) | −8.9 (−9.9 to −7.9) | 3.6 (1.8 to 5.5) |
| Any post-acute facility | 3.9 (1.9 to 5.8) | 2.9 (0.7 to 5.1) | −11.0 (−12.6 to −9.4) | 14.0 (11.2 to 16.7) |
| Whole-stay length of stay, days | 1.74 (1.12 to 2.35) | 1.60 (0.78 to 2.43) | 3.53 (2.77 to 4.30) | −1.93 (−3.06 to −0.81) |
| **C. Common period: Massachusetts 2017–2019 anchor year group vs Texas 2017–2019 (standardized logistic models)** |  |  |  |  |
| SNF or nursing facility, M0, admission-time | 5.2 (3.5 to 7.0) | — | 4.3 (3.8 to 4.8) | 0.9 (−0.9 to 2.8) |
| Inpatient rehabilitation facility, M0, admission-time | −1.6 (−3.4 to 0.1) | — | −2.6 (−2.9 to −2.4) | 1.1 (−0.7 to 2.8) |
| Long-term acute care hospital, M0, admission-time | 3.1 (1.8 to 4.5) | — | −1.3 (−1.4 to −1.2) | 4.4 (3.0 to 5.7) |
| Any post-acute facility, M0, admission-time | 6.7 (4.1 to 9.4) | — | 0.0 (−0.7 to 0.6) | 6.7 (4.0 to 9.5) |
| SNF or nursing facility, M3, full covariates | 4.5 (2.9 to 6.2) | — | 3.2 (2.7 to 3.6) | 1.4 (−0.3 to 3.1) |
| Inpatient rehabilitation facility, M3, full covariates | −1.4 (−3.1 to 0.2) | — | −2.7 (−3.0 to −2.4) | 1.3 (−0.4 to 3.0) |
| Long-term acute care hospital, M3, full covariates | 2.1 (1.0 to 3.3) | — | −1.6 (−1.7 to −1.4) | 3.7 (2.5 to 4.9) |
| Any post-acute facility, M3, full covariates | 5.3 (3.1 to 8.0) | — | −1.4 (−1.9 to −0.8) | 6.6 (4.1 to 9.1) |

Panels A and B: adjusted probability differences (Medicaid − private; for length of stay, differences in days) with 95% CIs from linear probability models; length of stay is the whole hospital stay in calendar days in both settings. Texas-comparable covariates are age group, sex, race and ethnicity, admission type, Charlson index category, invasive ventilation, and period; the Texas model includes hospital and quarter fixed effects and the full Texas covariate set. Panel C: standardized logistic models as in Table 2; Massachusetts restricted to the MIMIC-IV 2017–2019 anchor year group, which approximates admissions in those years. Last column: difference between the Massachusetts (panels A–B, Texas-comparable covariates) and Texas estimates; its standard error is the square root of the sum of the two squared standard errors, each recovered from its 95% CI as (upper − lower)/3.92, so Massachusetts robust or bootstrap uncertainty and Texas hospital clustering are both carried through and the samples are treated as independent. These differences describe the two data sources and do not isolate state policy.

**Table 4.**
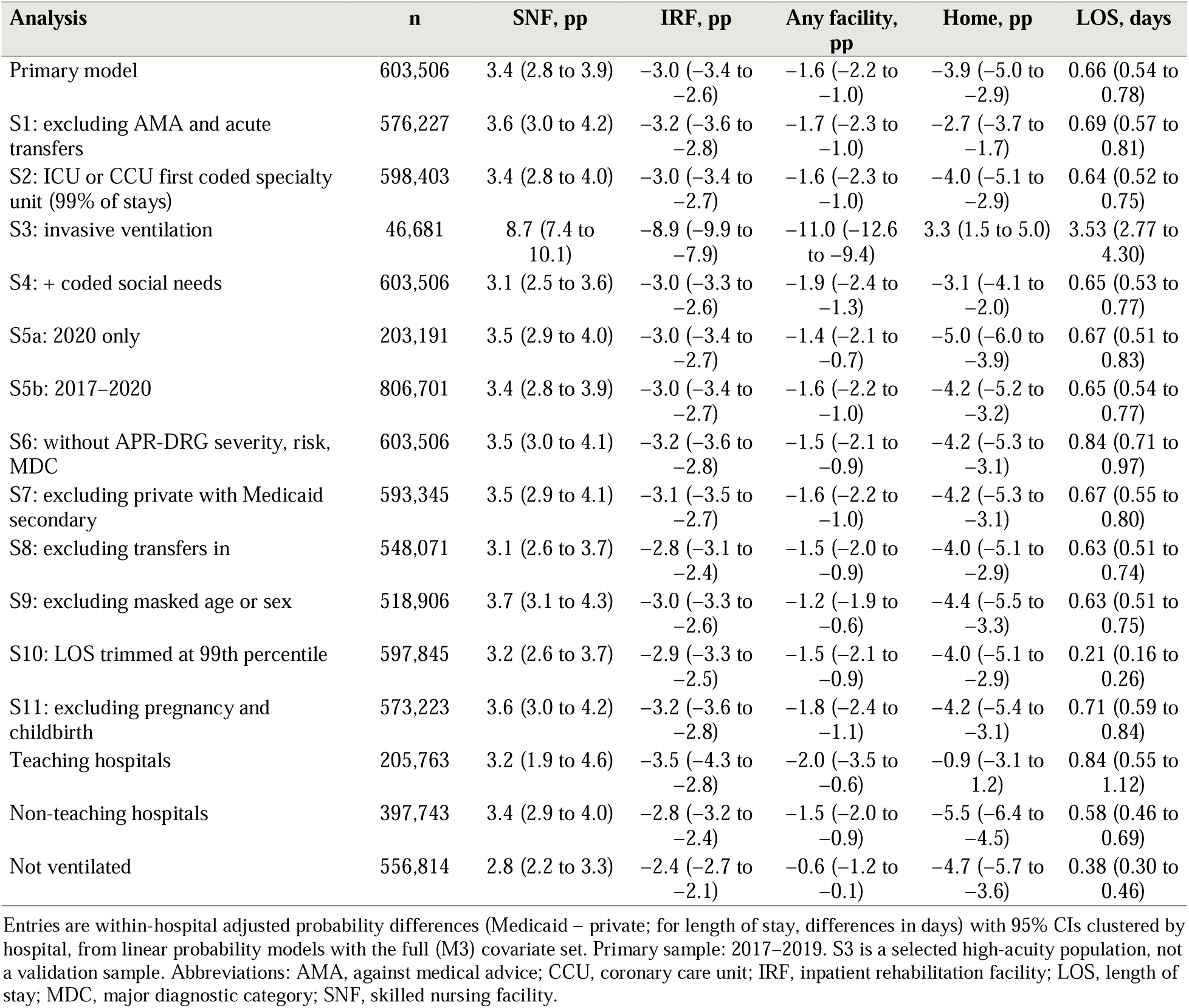
Texas sensitivity and subgroup analyses.

| **Analysis** | **n** | **SNF, pp** | **IRF, pp** | **Any facility, pp** | **Home, pp** | **LOS, days** |
|---|---|---|---|---|---|---|
| Primary model | 603,506 | 3.4 (2.8 to 3.9) | −3.0 (−3.4 to −2.6) | −1.6 (−2.2 to −1.0) | −3.9 (−5.0 to −2.9) | 0.66 (0.54 to 0.78) |
| S1: excluding AMA and acute transfers | 576,227 | 3.6 (3.0 to 4.2) | −3.2 (−3.6 to −2.8) | −1.7 (−2.3 to −1.0) | −2.7 (−3.7 to −1.7) | 0.69 (0.57 to 0.81) |
| S2: ICU or CCU first coded specialty unit (99% of stays) | 598,403 | 3.4 (2.8 to 4.0) | −3.0 (−3.4 to −2.7) | −1.6 (−2.3 to −1.0) | −4.0 (−5.1 to −2.9) | 0.64 (0.52 to 0.75) |
| S3: invasive ventilation | 46,681 | 8.7 (7.4 to 10.1) | −8.9 (−9.9 to −7.9) | −11.0 (−12.6 to −9.4) | 3.3 (1.5 to 5.0) | 3.53 (2.77 to 4.30) |
| S4: + coded social needs | 603,506 | 3.1 (2.5 to 3.6) | −3.0 (−3.3 to −2.6) | −1.9 (−2.4 to −1.3) | −3.1 (−4.1 to −2.0) | 0.65 (0.53 to 0.77) |
| S5a: 2020 only | 203,191 | 3.5 (2.9 to 4.0) | −3.0 (−3.4 to −2.7) | −1.4 (−2.1 to −0.7) | −5.0 (−6.0 to −3.9) | 0.67 (0.51 to 0.83) |
| S5b: 2017–2020 | 806,701 | 3.4 (2.8 to 3.9) | −3.0 (−3.4 to −2.7) | −1.6 (−2.2 to −1.0) | −4.2 (−5.2 to −3.2) | 0.65 (0.54 to 0.77) |
| S6: without APR-DRG severity, risk, MDC | 603,506 | 3.5 (3.0 to 4.1) | −3.2 (−3.6 to −2.8) | −1.5 (−2.1 to −0.9) | −4.2 (−5.3 to −3.1) | 0.84 (0.71 to 0.97) |
| S7: excluding private with Medicaid secondary | 593,345 | 3.5 (2.9 to 4.1) | −3.1 (−3.5 to −2.7) | −1.6 (−2.2 to −1.0) | −4.2 (−5.3 to −3.1) | 0.67 (0.55 to 0.80) |
| S8: excluding transfers in | 548,071 | 3.1 (2.6 to 3.7) | −2.8 (−3.1 to −2.4) | −1.5 (−2.0 to −0.9) | −4.0 (−5.1 to −2.9) | 0.63 (0.51 to 0.74) |
| S9: excluding masked age or sex | 518,906 | 3.7 (3.1 to 4.3) | −3.0 (−3.3 to −2.6) | −1.2 (−1.9 to −0.6) | −4.4 (−5.5 to −3.3) | 0.63 (0.51 to 0.75) |
| S10: LOS trimmed at 99th percentile | 597,845 | 3.2 (2.6 to 3.7) | −2.9 (−3.3 to −2.5) | −1.5 (−2.1 to −0.9) | −4.0 (−5.1 to −2.9) | 0.21 (0.16 to 0.26) |
| S11: excluding pregnancy and childbirth | 573,223 | 3.6 (3.0 to 4.2) | −3.2 (−3.6 to −2.8) | −1.8 (−2.4 to −1.1) | −4.2 (−5.4 to −3.1) | 0.71 (0.59 to 0.84) |
| Teaching hospitals | 205,763 | 3.2 (1.9 to 4.6) | −3.5 (−4.3 to −2.8) | −2.0 (−3.5 to −0.6) | −0.9 (−3.1 to 1.2) | 0.84 (0.55 to 1.12) |
| Non-teaching hospitals | 397,743 | 3.4 (2.9 to 4.0) | −2.8 (−3.2 to −2.4) | −1.5 (−2.0 to −0.9) | −5.5 (−6.4 to −4.5) | 0.58 (0.46 to 0.69) |
| Not ventilated | 556,814 | 2.8 (2.2 to 3.3) | −2.4 (−2.7 to −2.1) | −0.6 (−1.2 to −0.1) | −4.7 (−5.7 to −3.6) | 0.38 (0.30 to 0.46) |

Entries are within-hospital adjusted probability differences (Medicaid − private; for length of stay, differences in days) with 95% CIs clustered by hospital, from linear probability models with the full (M3) covariate set. Primary sample: 2017–2019. S3 is a selected high-acuity population, not a validation sample. Abbreviations: AMA, against medical advice; CCU, coronary care unit; IRF, inpatient rehabilitation facility; LOS, length of stay; MDC, major diagnostic category; SNF, skilled nursing facility.

**Table 5.** Sequential adjustment in temporal order: primary outcomes by model and setting.

| **Model** | **Mass.: SNF, pp** | **Mass.: IRF, pp** | **Mass.: LOS, days** | **Texas: SNF, pp** | **Texas: IRF, pp** | **Texas: LOS, days** |
|---|---|---|---|---|---|---|
| M0: admission-time characteristics (primary descriptive model) | 5.0 (4.2 to 5.8) | −2.0 (−2.8 to −1.2) | 1.91 (1.51 to 2.32) | 4.1 (3.5 to 4.7) | −2.6 (−3.0 to −2.3) | 1.41 (1.26 to 1.56) |
| M1: + Charlson index | 4.7 (4.0 to 5.5) | −2.2 (−3.0 to −1.4) | 1.59 (1.19 to 1.98) | 3.7 (3.1 to 4.3) | −3.1 (−3.5 to −2.7) | 1.06 (0.92 to 1.20) |
| M1p: M0 + Charlson from present-on-admission codes (Texas only) | — | — | — | 3.7 (3.1 to 4.3) | −3.0 (−3.4 to −2.6) | 1.11 (0.97 to 1.25) |
| M2: + organ support (and first-day SOFA in Massachusetts) | 4.5 (3.7 to 5.2) | −2.4 (−3.2 to −1.6) | 1.09 (0.72 to 1.47) | 3.5 (3.0 to 4.1) | −3.2 (−3.6 to −2.8) | 0.84 (0.71 to 0.97) |
| M3: + discharge severity, risk, MDC (Texas) or ICU course (Massachusetts) | 4.4 (3.7 to 5.2) | −1.9 (−2.7 to −1.1) | 1.29 (0.98 to 1.61) | 3.4 (2.8 to 3.9) | −3.0 (−3.4 to −2.6) | 0.66 (0.54 to 0.78) |
| M3p: M3 with present-on-admission Charlson (Texas only) | — | — | — | 3.4 (2.8 to 4.0) | −3.0 (−3.3 to −2.6) | 0.68 (0.56 to 0.80) |
| M4: + diagnosis-coded social-needs proxies | 4.0 (3.2 to 4.8) | −1.6 (−2.5 to −0.8) | 1.22 (0.89 to 1.56) | 3.1 (2.5 to 3.6) | −3.0 (−3.3 to −2.6) | 0.65 (0.53 to 0.77) |

Entries are adjusted probability differences (Medicaid − private, 95% CI) from linear probability models; length of stay is the difference in days from linear models. Covariates are added in temporal order (Figure 1; Supplementary Table S2). Massachusetts: heteroskedasticity-robust errors; M0 includes age, sex, race and ethnicity, admission type, and period; M2 adds first-day SOFA, ventilation, and vasopressors; M3 adds ICU length of stay and ICU type. Texas: hospital and quarter fixed effects with hospital-clustered errors; M0 includes age group, sex, race, ethnicity, admission type and source, and emergency department use; M2 adds ventilation; M3 adds APR-DRG severity, risk of mortality, and MDC. M1p and M3p replace the all-codes Charlson index with an index from present-on-admission codes (rare MDC levels pooled). The corresponding bounded estimates are in Supplementary Table S17.

## Discussion

In a single academic medical center in Massachusetts and 325 acute care hospitals in Texas, Medicaid, compared with private insurance, was associated with a similar pattern among working-age survivors of intensive care: more discharge to nursing facilities, less discharge to inpatient rehabilitation or home without services, and longer hospital stays. It was present with admission-time adjustment alone and, in Texas, within hospitals, in a bounded-probability model, across charge-defined subgroups, and without dependence on any single hospital. Destination differences changed modestly but persisted as covariates measured during the stay were added, whereas the LOS differences narrowed by a third to a half with adjustment for comorbidity and severity. Whether Medicaid patients received *more* facility-based care differed between settings, because LTCH use was higher with Medicaid in Massachusetts and lower in Texas.

The consistent finding — nursing facility rather than rehabilitation — extends results from working-age and registry stroke populations (Skolarus et al., 2012; Medford-Davis et al., 2016) to a broad, all-diagnosis critical illness population. Its appearance in two data sources, and within Texas hospitals, makes it unlikely to reflect one institution’s practices or the sorting of Medicaid patients to particular hospitals. It is compatible with the provider-acceptance, administrative, and social channels of the framework, which could operate in both states. The data cannot show whether the SNF placements were clinically appropriate, whether patients were eligible for or offered IRF care, or how patients fared after discharge.

The differences between settings are consistent with the benefit channel, but the design cannot attribute them to it. During the Texas study years, state regulation limited Medicaid payment for inpatient hospital care to the first 30 days of each spell of illness, with a new spell beginning only after 60 consecutive days out of an acute care facility (1 Tex. Admin. Code § 354.1072). The December 2018 and August 2019 editions of the state provider manual defined inpatient rehabilitation as a benefit when provided as part of a general acute care admission, or with prior authorization for clients aged 20 years or younger in a freestanding rehabilitation facility, made all rehabilitation subject to the spell-of-illness limit, and set a $200,000 annual inpatient limit for adults aged 21 years or older (Texas Medicaid & Healthcare Partnership, 2018, 2019; 1 Tex. Admin. Code § 354.1149). These rules did not apply uniformly: a state evaluation describes the spell-of-illness limit as applying to adults in fee-for-service Medicaid,

STAR+PLUS (the managed care program for adults with disabilities), and STAR Health, but not in STAR (the program for pregnant women and low-income parents), and the $200,000 limit as applying only in fee-for-service Medicaid and STAR Health (Texas Health and Human Services Commission, 2024). Texas was also among the states without Medicaid IRF coverage in 2010 (Skolarus et al., 2014). We could not obtain a 2017 manual, the manual does not address LTCH stays, and the payer field does not identify program, so which stays were subject to which rule is unknown. The very low Texas ratios for IRF and LTCH discharge, their concentration among ventilated patients, and the relative increases in SNF and home health discharge are consistent with these rules. In Massachusetts, whose Medicaid regulations provide for chronic disease and rehabilitation hospital services (MassHealth, 2015), the IRF difference was smaller and LTCH use was higher with Medicaid; the regulation does not show that all enrollees had equivalent access to these settings. The Texas LTCH finding also matches evidence from Pennsylvania (Lane-Fall et al., 2012), where these rules did not apply. The contrast therefore motivates a hypothesis — that adult Medicaid benefit design shapes the type of post-acute care after critical illness — that requires designs using policy variation to test.

Eligibility composition also differs. During the study period, Texas Medicaid covered parents only up to 17% of the federal poverty level and did not cover other nondisabled adults, whereas Massachusetts covered adults up to 138% (Kaiser Family Foundation & Georgetown University Center for Children and Families, 2019). Working-age Texas Medicaid patients are therefore more likely to qualify through disability, pregnancy, or extreme poverty, and may have functional and social needs that administrative data do not capture. This could contribute to the larger Texas differences, although greater need alone would be expected to increase, not decrease, IRF and LTCH use, and excluding pregnancy stays did not change the results.

Longer stays with Medicaid in both settings, concentrated among patients discharged to facilities, are compatible with documented delays in authorization and placement (Sorensen et al., 2020; Haddad et al., 2025) and with Massachusetts reports of payer-related transition delays (Massachusetts Health & Hospital Association, 2025; Massachusetts TACPAC Task Force, 2025), but LOS also reflects severity, complications, and the ICU course — adjustment for these narrowed the differences — and neither data source records when patients were medically ready for discharge.

### Policy implications

The findings have three implications for research and practice, none of which assumes that a shift between settings is harmful. First, studies of post-acute access should report destinations separately; in Texas, a single facility composite showed little difference by payer while concealing large, offsetting SNF, IRF, and LTCH differences. Second, adult Medicaid coverage of IRF and LTCH care and inpatient day limits are explicit, documented benefit choices that are consistent with the largest differences here; they merit direct evaluation with designs that exploit changes in these rules, linked to clinical eligibility, program enrollment, and post-discharge outcomes. Third, hospitals and Medicaid plans could track destination and LOS by payer alongside referral, authorization, and acceptance data to identify which channel operates.

### Limitations

This study has limitations. The settings are not equivalent in unit of analysis, hospitals, covariates, or measurement, and the Massachusetts common-period subset only approximates 2017–2019, so differences between them cannot be attributed to state policy. Insurance was measured from administrative records, which may misclassify coverage, program, plan type, and dual eligibility; in Texas, some Medicaid managed care claims may be coded as commercial. Benefit rules could not be linked to individual stays. The Texas cohort is defined by billing categories that include intermediate, burn, trauma, and psychiatric intensive care charges, and repeated stays by the same patient cannot be identified, so hospital-clustered errors do not capture dependence among stays by one patient at different hospitals. Discharge destination fields can misclassify LTCH transfers (Kahn & Iwashyna, 2010), and destinations are harmonized administrative categories. Covariates measured after admission may be affected by care processes, and Massachusetts comorbidity codes lack present-on-admission indicators. Administrative data lack pre-admission function, caregiver support, and preferences, and the diagnosis-coded social-needs proxies may be incompletely and differentially coded. We did not observe whether IRF or LTCH care was requested, denied, or declined, or any post-discharge outcomes. Finally, Setting 1 is a single academic center.

## Conclusion

Among working-age survivors of intensive care at a Massachusetts academic medical center and Texas hospitalizations with ICU or coronary care unit accommodation charges, Medicaid was consistently associated with nursing facility rather than rehabilitation discharge and with longer hospital stays, whereas overall facility use differed between settings. Texas Medicaid rules limiting adult freestanding rehabilitation and, in some programs, inpatient days are consistent with the Texas pattern and warrant direct evaluation.

## Data availability

MIMIC-IV is available to credentialed users through PhysioNet. The Texas Inpatient Public Use Data File is available from the Texas Department of State Health Services under a data use agreement. Row-level data cannot be shared by the author. Analysis code (SQL and Python, without data or outputs) is available at https://github.com/nasiruddinstudents-ctrl/insurance-post-icu-transitions (release v1.0, commit 062d2a3).

## Ethics

MIMIC-IV was approved by the institutional review boards of Beth Israel Deaconess Medical Center and the Massachusetts Institute of Technology, with a waiver of informed consent; data were accessed through PhysioNet under a credentialed data use agreement. The Texas Hospital Inpatient Discharge Public Use Data File is a de-identified public use file released by the Texas Department of State Health Services under a data use agreement. The study used only de-identified data. The analyses, interpretations, and conclusions are the author’s and not those of the Texas Department of State Health Services.

## Author contributions

M.N.U. is the sole author and conceived and designed the study, analyzed the data, interpreted the results, and wrote the manuscript.

## Use of generative artificial intelligence

Generative AI assistants were used to help draft analysis code and manuscript text (Claude, Anthropic) and to support the literature search (Elicit). The author ran all analyses, verified all code, results, and text, and takes full responsibility for the content. No patient-level data were provided to any AI tool.

## References

1 Tex. Admin. Code § 354.1072 (Authorized inpatient hospital services). Texas Health and Human Services Commission. https://regulations.justia.com/states/texas/title-1/part-15/chapter-354/subchapter-a/division-6/section-354-1072

1 Tex. Admin. Code § 354.1149 (Benefits and limitations). Texas Health and Human Services Commission. https://regulations.justia.com/states/texas/title-1/part-15/chapter-354/subchapter-a/division-11/section-354-1149

Andersen, R. M. (1995). Revisiting the behavioral model and access to medical care: Does it matter? Journal of Health and Social Behavior, 36(1), 1–10. 10.2307/2137284

Austin, P. C., & Stuart, E. A. (2015). Moving towards best practice when using inverse probability of treatment weighting (IPTW) using the propensity score to estimate causal treatment effects in observational studies. Statistics in Medicine, 34(28), 3661–3679. 10.1002/sim.6607

Benchimol, E. I., Smeeth, L., Guttmann, A., Harron, K., Moher, D., Petersen, I., et al. (2015). The REporting of studies Conducted using Observational Routinely-collected health Data (RECORD) statement. PLoS Medicine, 12(10), e1001885. 10.1371/journal.pmed.1001885

Charlson, M. E., Pompei, P., Ales, K. L., & MacKenzie, C. R. (1987). A new method of classifying prognostic comorbidity in longitudinal studies: Development and validation. Journal of Chronic Diseases, 40(5), 373–383. 10.1016/0021-9681(87)90171-8

Deb, P., & Norton, E. C. (2018). Modeling health care expenditures and use. Annual Review of Public Health, 39, 489–505. 10.1146/annurev-publhealth-040617-013517

Haddad, D. N., Hatchimonji, J. S., Eisinger, E. C., Chen, A. T., Ramadan, O. I., Morgan, A. U., et al. (2025). Awaiting insurance coverage: Medicaid enrollment and post-acute care use after traumatic injury. Journal of Trauma and Acute Care Surgery, 98(3), 418–424. 10.1097/TA.0000000000004550

Johnson, A. E. W., Bulgarelli, L., Shen, L., Gayles, A., Shammout, A., Horng, S., et al. (2023). MIMIC-IV, a freely accessible electronic health record dataset. Scientific Data, 10, 1. 10.1038/s41597-022-01899-x

Johnson, A. E. W., Stone, D. J., Celi, L. A., & Pollard, T. J. (2018). The MIMIC Code Repository: Enabling reproducibility in critical care research. Journal of the American Medical Informatics Association, 25(1), 32–39. 10.1093/jamia/ocx084

Johnson, A., Bulgarelli, L., Pollard, T., Gow, B., Moody, B., Horng, S., et al. (2024). *MIMIC-IV* (version 3.1) [Data set]. PhysioNet. 10.13026/kpb9-mt58

Kahn, J. M., Benson, N. M., Appleby, D., Carson, S. S., & Iwashyna, T. J. (2010). Long-term acute care hospital utilization after critical illness. JAMA, 303(22), 2253–2259. 10.1001/jama.2010.761

Kahn, J. M., & Iwashyna, T. J. (2010). Accuracy of the discharge destination field in administrative data for identifying transfer to a long-term acute care hospital. BMC Research Notes, 3, 205. 10.1186/1756-0500-3-205

Kaiser Family Foundation & Georgetown University Center for Children and Families. (2019). Medicaid and CHIP eligibility, enrollment, and cost sharing policies as of January 2019: Findings from a 50-state survey (Table 5: Medicaid income eligibility limits for adults). https://files.kff.org/attachment/Report-Medicaid-and-CHIP-Eligibility-as-of-Jan-2019-Table-5

Kane, W. G., Wright, D. A., Fu, R., & Carlson, K. F. (2014). Racial/ethnic and insurance status disparities in discharge to posthospitalization care for patients with traumatic brain injury. Journal of Head Trauma Rehabilitation, 29(6), E10–E17. 10.1097/HTR.0000000000000028

Lane-Fall, M. B., Iwashyna, T. J., Cooke, C. R., Benson, N. M., & Kahn, J. M. (2012). Insurance and racial differences in long-term acute care utilization after critical illness. Critical Care Medicine, 40(4), 1143–1149. 10.1097/CCM.0b013e318237706b

Massachusetts Health & Hospital Association. (2025). The case for post-acute care reforms in Massachusetts. https://www.mhalink.org/voicesinhealthcare/the-case-for-post-acute-care-reforms-in-massachusetts/

Massachusetts Transitions from Acute Care to Post-Acute Care (TACPAC) Task Force. (2025). Report to the Legislature. Executive Office of Health and Human Services. https://www.mass.gov/doc/tacpac-task-force-report-final-7-31-2025-0/download

MassHealth. (2015). 130 CMR 435.000: Chronic disease and rehabilitation inpatient hospital services. Commonwealth of Massachusetts, Executive Office of Health and Human Services. https://www.mass.gov/regulations/130-CMR-43500-chronic-disease-and-rehabilitation-inpatient-hospital-services

Medford-Davis, L. N., Fonarow, G. C., Bhatt, D. L., Xu, H., Smith, E. E., Suter, R., et al. (2016). Impact of insurance status on outcomes and use of rehabilitation services in acute ischemic stroke: Findings from Get With The Guidelines-Stroke. Journal of the American Heart Association, 5(11), e004282. 10.1161/JAHA.116.004282

Muller, C. J., & MacLehose, R. F. (2014). Estimating predicted probabilities from logistic regression: Different methods correspond to different target populations. International Journal of Epidemiology, 43(3), 962–970. 10.1093/ije/dyu029

Needham, D. M., Davidson, J., Cohen, H., Hopkins, R. O., Weinert, C., Wunsch, H., et al. (2012). Improving long-term outcomes after discharge from intensive care unit: Report from a stakeholders’ conference. Critical Care Medicine, 40(2), 502–509. 10.1097/CCM.0b013e318232da75

Pollard, T., Moody, B. E., Lehman, L., Gow, B., Fernandes, C., Xie, C., et al. (2026). PhysioNet as a global platform for biomedical research. Nature Health. 10.1038/s44360-026-00096-z

Quan, H., Sundararajan, V., Halfon, P., Fong, A., Burnand, B., Luthi, J.-C., et al. (2005). Coding algorithms for defining comorbidities in ICD-9-CM and ICD-10 administrative data. Medical Care, 43(11), 1130–1139. 10.1097/01.mlr.0000182534.19832.83

Shah, R. (2025). Impact of insurance payor on ICU length of stay among stroke patients: A retrospective cohort study using MIMIC-IV [Abstract 4365176]. Circulation, 152(Suppl. 3). 10.1161/circ.152.suppl_3.4365176

Shah, R., & Strobel, S. (2026). Association of insurance payor and discharge to inpatient rehabilitation among ICU ischemic stroke patients receiving tPA [Abstract]. Neurology. American Academy of Neurology Annual Meeting. 10.1212/WNL.0000000000216956

Skolarus, L. E., Burke, J. F., Morgenstern, L. B., Meurer, W. J., Adelman, E. E., Kerber, K. A., et al. (2014). Impact of state Medicaid coverage on utilization of inpatient rehabilitation facilities among patients with stroke. Stroke, 45(8), 2472–2474. 10.1161/STROKEAHA.114.005882

Skolarus, L. E., Meurer, W. J., Burke, J. F., Prvu Bettger, J., & Lisabeth, L. D. (2012). Effect of insurance status on postacute care among working age stroke survivors. Neurology, 78(20), 1590–1595. 10.1212/WNL.0b013e3182563bf5

Sorensen, M., Sercy, E., Salottolo, K., Waxman, M., West, T. A., Tanner, A., et al. (2020). The effect of discharge destination and primary insurance provider on hospital discharge delays among patients with traumatic brain injury: A multicenter study of 1,543 patients. Patient Safety in Surgery, 14, 2. 10.1186/s13037-019-0227-z

Texas Department of State Health Services, Center for Health Statistics. (2018–2021). Texas Hospital Inpatient Discharge Public Use Data File, first quarter 2017 through fourth quarter 2020 [Data set]. Austin, TX: Texas Department of State Health Services.

Texas Department of State Health Services, Texas Health Care Information Collection. (2020). Texas Hospital Inpatient Discharge Public Use Data File user manual, 2019 (Document E25-14163). Austin, TX: Texas Department of State Health Services.

Texas Health and Human Services Commission. (2024). COVID-19 1115(a) demonstration evaluation: Inpatient hospitalization limitations (Final report). Office of Data, Analytics, and Performance. https://www.medicaid.gov/medicaid/section-1115-demonstrations/downloads/tx-hlth-trnsfrmn-cvid19-115a-dmnst-fnl-rprt-05102024-updated.pdf

Texas Medicaid & Healthcare Partnership. (2018). Texas Medicaid provider procedures manual, Vol. 2: Inpatient and outpatient hospital services handbook (December 2018; Sections 3.1.1, 3.1.2, and 3.3). Texas Health and Human Services Commission. https://www.tmhp.com/sites/default/files/file-library/resources/provider-manuals/tmppm/pdf-chapters/2018/2018-12-2_Inpatient_Outpatient_Hosp_Srvs.pdf

Texas Medicaid & Healthcare Partnership. (2019). Texas Medicaid provider procedures manual, Vol. 2: Inpatient and outpatient hospital services handbook (August 2019; Section 3.3). Texas Health and Human Services Commission. https://www.tmhp.com/sites/default/files/file-library/resources/provider-manuals/tmppm/pdf-chapters/2019/2019-08-2_Inpatient_Outpatient_Hosp_Srvs.pdf

VanderWeele, T. J., & Ding, P. (2017). Sensitivity analysis in observational research: Introducing the E-value. Annals of Internal Medicine, 167(4), 268–274. 10.7326/M16-2607

Vincent, J.-L., Moreno, R., Takala, J., Willatts, S., De Mendonça, A., Bruining, H., et al. (1996). The SOFA (Sepsis-related Organ Failure Assessment) score to describe organ dysfunction/failure. Intensive Care Medicine, 22(7), 707–710. 10.1007/BF01709751

von Elm, E., Altman, D. G., Egger, M., Pocock, S. J., Gøtzsche, P. C., Vandenbroucke, J. P., & STROBE Initiative. (2007). The Strengthening the Reporting of Observational Studies in Epidemiology (STROBE) statement: Guidelines for reporting observational studies. The Lancet, 370(9596), 1453–1457. 10.1016/S0140-6736(07)61602-X

Zogg, C. K., Scott, J. W., Metcalfe, D., Gluck, A. R., Curfman, G. D., Davis, K. A., et al. (2019). Association of Medicaid expansion with access to rehabilitative care in adult trauma patients. JAMA Surgery, 154(5), 402–411. 10.1001/jamasurg.2018.5177

